# Resource need and cost estimates for universal health coverage across 122 countries using the WHO UHC Compendium

**DOI:** 10.64898/2026.03.12.26347243

**Authors:** Charlie Nederpelt, Gerard Abou Jaoude, Gavin Surgey, Baktygul Isaeva, Saltanat Zhetibaeva, Han Win Htat, Hassan Haghparast-Bidgoli, Rob Baltussen

## Abstract

**Introduction:** Reliable estimates of resource requirements for essential health interventions are critical for universal health coverage (UHC) planning. Existing benchmarks provide limited operational guidance for national decision-making.

**Methods:** We developed an ingredient-based costing model aligned with the WHO Universal Health Coverage Compendium (UHCC), which specifies delivery platforms, actions and technologies for 544 interventions. We estimated resource needs and costs associated with delivery of all UHCC-defined interventions at 80% coverage across 122 low- and middle-income countries, applying a multimorbidity adjustment to reduce potential double counting and using authoritative epidemiological, demographic and cost data.

**Results:** Modeled delivery of Core UHCC interventions is estimated at USD 2.0 trillion annually (5.7% of aggregate gross domestic product) or USD$249, 294 and 363 per capita in low-, lower-middle- and upper-middle-income countries, respectively. Cost estimates closely aligned with WHO projections for achieving Sustainable Development Goal 3, but were 1.7-2.7x higher than Disease Control Priorities Network internal cost estimates.

**Conclusion:** The UHCC aligned cost model provides transparent resource need and cost data under normative service delivery assumptions, and offers a practical starting point for country-level contextualization for health service packages planning.

## Introduction

Countries use Essential Packages of Health Services (EPHS) to define which interventions should be provided, for whom, and with what level of financial coverage.(1) EPHS, or packages, are central policy instruments for making explicit priorities on the path to UHC.(2,3) Poorly designed packages can result in implementation failures, including entitlements that exceed fiscal capacity, diversion of resources away from high-value care, or vague intervention definitions that hinder operationalization.(4) By contrast, well-specified EPHS, combined with political commitment and adequate financing, support effective service delivery and system resilience, including during major shocks such as the COVID-19 pandemic and reductions in external health financing.(5,6)

Robust costing evidence is essential for both EPHS design and implementation, underpinning budgeting and strategic purchasing decisions.(7–10) However, producing reliable cost and resource estimates remains technically demanding and data-intensive, and national capacity for such analyses is limited in many low- and middle-income countries (LMICs).(11,12)

While international guidance exists, most tools focus on individual interventions rather than entire health-system packages.(13–15) To our knowledge, only two tools have previously estimated EPHS costs across the entire health system: the WHO model for Sustainable Development Goal 3 which costed 189 interventions, and the Disease Control Priorities (DCP) network’s estimate for its Essential Universal Health Coverage (EUHC) and Highest Priority Package (HPP), covering 218 and 108 UHC-relevant interventions, respectively. (16,17)

In this paper, we present the global application of a new costing model built using the data and structure of available in the WHO Universal Health Coverage Compendium (UHCC), which draws directly on WHO expert input to support EPHS design and implementation.(18) The UHCC does not constitute a WHO-recommended or minimum UHC package but rather provides a list of interventions to be considered for integrated service delivery, by detailing specific actions and inputs, and explicitly incorporating foundational care, referral pathways, and delivery platforms. This level of detail enables system-wide cost estimation for implementable interventions and packages.

We estimated detailed resource needs and costs of delivering 544 UHCC-defined interventions across 122 LMICs, using a purpose-built costing tool. The cost estimates presented here quantify the resources required to deliver these interventions as specified. Our aim was to provide transparent estimates of resource requirements and support country-level contextualization and planning of EPHS.

## Methods

An MS Excel-based (2021, version 2512) model was developed to estimate annual recurring costs for health service delivery from the provider perspective. Unit costs were collected in the most recent available cost year, with original currency and year reported, and converted to USD using the 2025 midyear conversion rate.(19) A total of 122 countries were included for model development based on availability of: i) Gross Domestic Product (GDP) data, ii) United Nations (UN) population estimates, and iii) Institute for Health Metrics and Evaluation (IHME) 2023 Global Burden of Disease study (GBD) incidence and prevalence rates.(20,21) Together, these countries represented approximately 100% of both LMIC population and GDP.

The July 1, 2024 UHCC version was used. The UHCC distinguishes between Core and Additional interventions. “Core” interventions as specified in the UHCC represent the baseline set of clinically effective interventions for a certain condition, that are feasible and applicable across contexts. “Additional” interventions reflect supplementary diagnostic or therapeutic options with higher resource intensity. In this study, Complete UHCC interventions refer to the combined set of Core and Additional interventions.

For each intervention, the UHCC normatively defines interventions, including i) description of the population in need (PiN), and the proportion of the PiN per encounter, ii) encounter type and frequency, iii) health worker cadres involved and time for each encounter, iv) diagnostic tests, v) medications and dosing regimens, vi) procedures, vii) required equipment, medical technologies and consumables, viii) and the distribution of encounters across delivery platforms. The complete model structure is explained in the **Supplement**. Frequency ranges (e.g. 2-4 visits, 4-6 weeks dosing) were replaced by the range mean. In <2% of encounters, missing encounter frequencies, dosing regimen and/or PiN shares were inferred using the intervention context, and with reference to treatment guidelines.

Ingredient-based costing was used for healthcare worker labor, drugs, in vitro diagnostics (IVDs), radiological exams (including equipment amortization) and durable medical products. To reduce the risk of double counting labor inputs for patients with multiple chronic conditions, outpatient labor costs were adjusted at package-level using a multimorbidity factor equal to half the ratio between pooled estimates of chronic multimorbidity prevalence (22-37%) and overall morbidity prevalence in LMICs (54%).(22,23) This adjustment was applied to high-prevalence, co-existing diseases based on diagnosis cluster analysis from primary care, i.e. cardiometabolic disease, osteoarthritis, chronic liver disease and viral hepatitis, and vision and hearing impairment.(24)

Single-use consumables (e.g. gloves, paper) and low-cost reusable items (e.g. stethoscopes) were costed as a percentage mark-up to labor, drug and tests, differentiated by encounter type and delivery platform. Facility-level overhead costs including administrative overhead, real estate maintenance and amortization, information technology, security etc., were costed as a percentage mark-up to labor, drug and tests, consistent with reported ranges from LMIC costing studies (10-30% overhead for community and outpatient care; 17-35% for hospital-based care).(25–29) System-level overheads such as governance were excluded.

Populations-in-need (PiN) were estimated for all 2,050 encounters in each country using a hierarchical approach. Where available, population shares specified directly in the UHCC were applied (13% of all UHCC encounters). Otherwise, country-specific demographic or cause-specific estimates from UN agencies were used (7%), followed by country-specific IHME GBD incidence and prevalence estimates combined with UN population data (68%). Where neither source was available, pooled estimates from peer-reviewed international meta-analyses were applied, prioritizing subregional over global estimates (17%) (**Table S1**).(27,28) For each country and intervention, unit costs were multiplied by the estimated PiN and by a target coverage of 80% across all interventions, consistent with DCP-3 costing.(17) This avoids introducing condition-specific prioritization bias.

Annual total and per capita resource need and costs were estimated for each country and compared across World Bank income groups (using 2025 fiscal year thresholds). To facilitate comparison with earlier global EPHS costing exercises, resource needs and costs for Core UHCC interventions were compared to personal interventions included in the DCP3 EUHC package, as mapped in the UHCC digital interface: the Service Package Design and Implementation tool.(30) To assess external validity and model robustness, we compared estimated resource need and costs with results from authoritative scientific literature and United Nations system reports.

We performed one-way sensitivity analyses on per capita cost per component, and on per capita cost across components for varying PiNs. Relative variations of +/-25% were applied to wages, IVDs and products. We modeled minimum and maximum overhead mark-up at 10-30% for primary care and 17-35% for hospital care, based on LMIC costing studies.(25–29) For medication, we applied relative variations of +/-33, 50 and 75% for drugs with unit costs of <$0.10, $0.10-10.00 and >$10.00 per daily defined dose, based on analysis of reported buyers price variation.(14) For PiNs, we applied relative variation of +/- 2% for UN population estimates, modeled the reported upper and lower bound for PiNs sourced from GBD incidence and prevalence rates, and applied +/- 20% for PiNs estimated using literature sources, roughly matching the average ratio of upper and lower bounds to point estimate values for GBD incidence and prevalence rates.(20,21)

## Results

### Total and per capita costs

The cost of delivering Complete UHCC interventions at 80% coverage across included LMICs in 2025 is estimated at USD 2.5 trillion (2025 prices), corresponding to 7.1% of aggregate GDP (**Table 1**). By comparison, Core UHCC interventions cost USD 2.0 trillion, or 5.7% of GDP. Population-weighted per capita costs of Core UHCC interventions are estimated at USD 249 in low-income countries (LICs), USD 294 in lower-middle-income countries (LMICs) and USD 363 in upper-middle-income countries (UMICs), corresponding to GDP shares of 35%, 12% and 4.1%, respectively. Per capita intervention costs ranged from (rounded) $0.00 for treatment of radiation and nuclear exposure (across income groups) to $22 per capita for “additional management of overweight and obesity with medications” in LIC and LMIC, and $42/capita for “diagnosis and treatment of stroke with medication” in UMIC. A full list of population-weighted per capita cost estimates is included in **Supplement 2**.

**Table 1.**
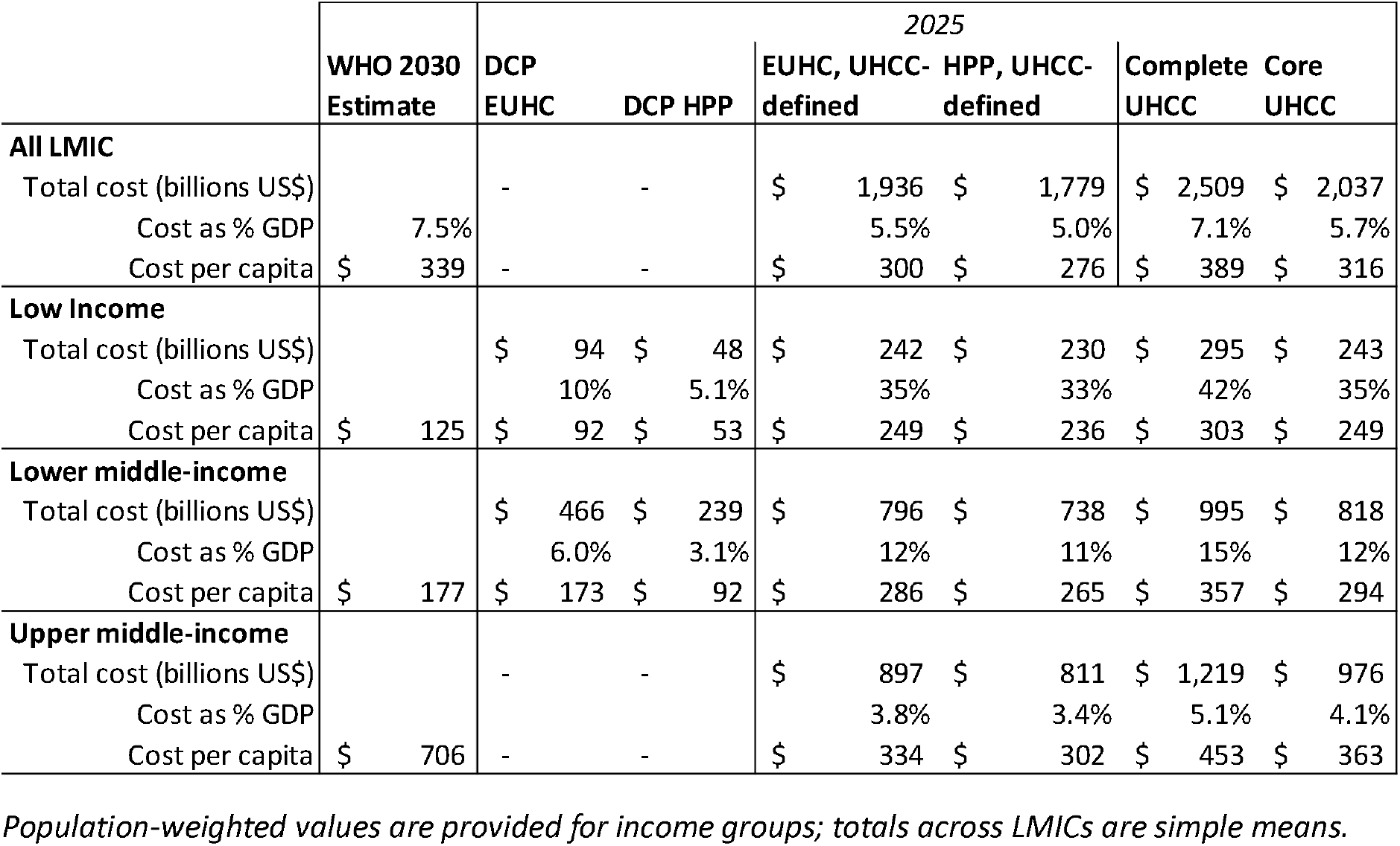
Comparison of total cost, cost as share of GDP and cost per capita for selected packages of interventions for achieving universal health coverage across World Bank income groups. Costs in inflation-adjusted 2025 US$.

When DCP-3 EUHC interventions are costed using UHCC intervention definitions, they account for 95% of Core UHCC costs and 77% of Complete UHCC costs, indicating substantial overlap. Core UHCC cost estimates are nevertheless 1.7–2.7 times higher than DCP-3’s internal EUHC estimates in LICs and LMICs, reflecting differences in intervention specification. Core UHCC intervention per capita cost estimates constitute 93% of WHO projections for achieving SDG3 ($316 vs $339 per capita across LMICs), although estimates diverge across income groups.

### Resource needs and utilization

Delivering Complete UHCC interventions at 80% coverage would require an estimated 615 full-time equivalents (FTE) of labor per 100,000 population (**Table 2**). Estimated utilization amounts to 23 outpatient visits and 2.7 inpatient admission days per person per year (p.p.p.y.). Procedure utilization is estimated at 1.0 procedure p.p.p.y., with radiology utilization of 0.16 X-rays, 0.18 ultrasounds, 0.003 MRI scans and 0.03 CT scans p.p.p.y. Utilization of in vitro diagnostics is estimated at 9.9 tests p.p.p.y. Medication consumption corresponds to 24 antibiotic, 8 antiretroviral, 0.4 antimalarial, 37 hypoglycaemic and 68 antihypertensive defined daily doses (DDD) p.p.p.y. These utilization estimates reflect normative models of care specified across all UHCC interventions.

**Table 2.** Sum of resources used in UHC Compendium interventions and UHCC defined DCP-3 service packages, total and population-weighted per capita average across 122 countries.

| Factor | Complete UHCC |  | Core UHCC services |  | DCP EUHC* |  | DCP HPP* |  |
| --- | --- | --- | --- | --- | --- | --- | --- | --- |
|  | Total<br>(n millions) | Per 100,000<br>population | Total<br>(n millions) | Per 100,000<br>population | Total<br>(n millions) | Per 100,000<br>population | Total<br>(n millions) | Per 100,000<br>population |
| Population | 6,448 |  |  |  |  |  |  |  |
| Labor (fte) | 40 | 615 | 37 | 573 | 33 | 506 | 29 | 444 |
| Outpatient encounters (fte) | 23 | 355 | 22 | 347 | 18 | 274 | 15 | 234 |
| Inpatient admissions (fte) | 8.6 | 134 | 7.3 | 113 | 7.9 | 122 | 7.3 | 114 |
| Procedures (excl. Radiology, fte) | 7.8 | 120 | 7.0 | 109 | 6.8 | 105 | 6.0 | 93 |
| Radiology examinations (fte) | 0.4 | 5.5 | 0.3 | 4.0 | 0.3 | 5.0 | 0.3 | 4.3 |
| Outpatient encounters | 148,628 | 2,311,537 | 144,800 | 2,252,796 | 113,170 | 1,780,007 | 97,769 | 1,544,576 |
| Inpatient admissions (in days) | 17,154.5 | 267,705 | 14,508.9 | 229,378 | 15,635.7 | 244,793 | 14,566.5 | 229,538 |
| Procedures (excl. Radiology) | 6,572 | 101,676 | 6,148 | 94,776 | 5,239 | 82,845 | 4,473 | 71,057 |
| X-ray exams | 1,063 | 16,445 | 1,039 | 16,044 | 1,024 | 15,841 | 636 | 9,895 |
| Ultrasounds | 1,171 | 18,486 | 747.4 | 11,806 | 1,112.5 | 17,627 | 1,112.4 | 17,625 |
| MRI scans | 21 | 314 | 5.3 | 74 | 7.8 | 118 | 7.7 | 117 |
| CT scans | 216 | 3,121 | 108 | 1,534 | 191 | 2,779 | 191 | 2,773 |
| Laboratory tests | 61,356 | 988,662 | 60,685 | 978,910 | 57,695 | 935,687 | 54,635 | 892,747 |
| Antibiotic doses (DDD) | 158,631 | 2,368,574 | 157,830 | 2,354,793 | 77,200 | 1,253,193 | 77,033 | 1,250,889 |
| ART doses (DDD) | 43,623 | 807,706 | 43,623 | 807,706 | 43,623 | 807,706 | 43,623 | 807,706 |
| Antimalarial doses (DDD) | 1,516 | 41,327 | 1,516 | 41,327 | 1,516 | 41,327 | 1,516 | 41,327 |
| Hypoglycaemics (DDD) | 271,438 | 3,774,914 | 266,425 | 3,700,376 | 266,425 | 3,700,375 | 266,425 | 3,700,375 |
| Antihypertensives (DDD) | 455,903 | 6,770,550 | 455,903 | 6,770,550 | 179,434 | 2,632,459 | 171,456 | 2515476 |

Total counts in x1,000,000, unless specified otherwise.
\*Service delivery for these packages is modeled using the UHCC definitions.
fte: full-time equivalent; MRI: magnetic resonance imaging; CT: computerized tomography; DDD: daily defined dose; ART: antiretroviral, DCP: disease control priorities; EUHC: essential universal health coverage; HPP: highest priority package

Within the UHCC, inclusion of Additional interventions is associated with modest increases in labor and laboratory inputs, but significantly higher utilization of e.g. MRI and CT scans. When analyses are restricted to clinical interventions included in both the UHCC and DCP frameworks, Complete and Core UHCC packages require higher resource utilization than both the DCP-3 EUHC and its more restricted HPP package.

### Cost composition by component, disease area and delivery platform

**Figure 1** presents per capita costs disaggregated by cost component and intervention category. Non-communicable diseases account for a larger share of costs than communicable diseases across all income groups, with the share attributable to non-communicable diseases increasing with country income level. The breakdown by cost components identified medicine ($1,002 billion, 40%) medical products ($423 billion, 17%) and overhead ($569 billion, 23%) as the largest cost drivers across LMICs, with smaller shares for health worker costs ($376 billion, 15%) and IVDs ($139 billion, 6%). By delivery platform, community-based encounters account for 4% of total costs, outpatient clinics for 22%, first-level referral hospitals for 36%, and second-level referral hospitals for 39% of total costs (**Figure S2**).

**Figure 1.**
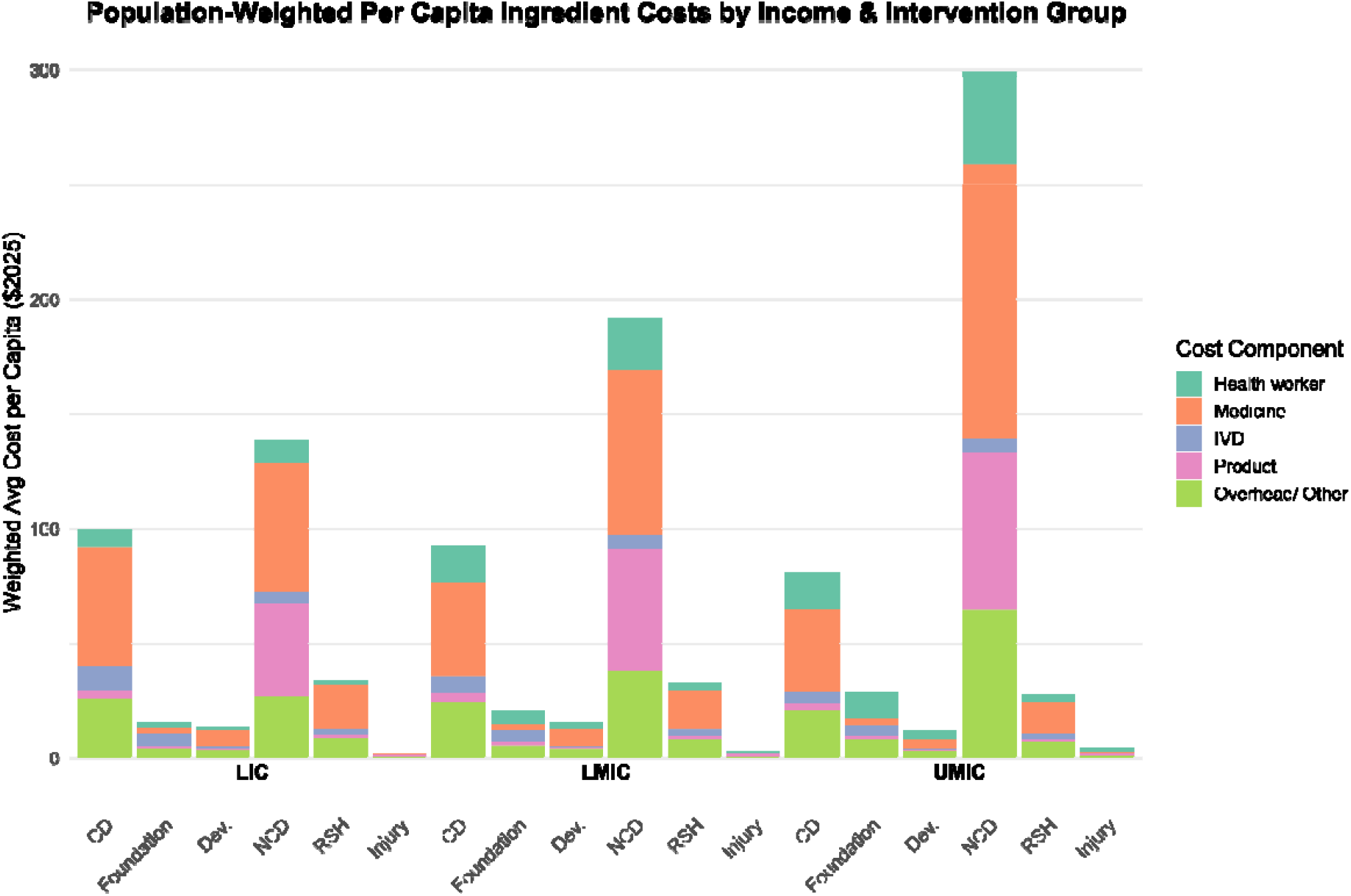
Population-weighted per capita cost of ingredients within intervention groups and World Bank Income Groups Left to right: CD: communicable disease; Foundations of care; Dev.: Growth, development and ageing; NCD: non-communicable disease and mental health; RSH: reproductive and sexual health; Injury and Violence

### Model robustness and external validity

Detailed results from robustness and external validity analyses are presented in **Supplement 3**. In brief, estimated human resource requirements of 6.15 FTE per 1,000 population were 18-43% lower than projections reported by WHO (7.6 and 9.0/1,000 population for LIC and MIC) and IHME (10.9/1,000 population across LMICs).(31,32) Outpatient and inpatient utilization rates exceed those reported in previous global estimates, reflecting the UHCC’s inclusion of high-frequency preventive interventions and chronic care management, including those not routinely provided at scale in high-income countries. Radiological procedures and laboratory testing utilization fall within reported ranges. Across key medication categories, normative utilization rates based on UHCC care models are 3, 4 and 9 times higher than reported current use of antihypertensives, antibiotics and hypoglycemics. Estimated normative per case costs for diagnosis and/or treatment of tuberculosis and malaria were generally comparable to published literature, with estimated HIV treatment cost ca. 20% lower in UHCC.

### Sensitivity analysis

One-way sensitivity analyses illustrate uncertainty around cost estimates (**Supplementary table S4**). Expected uncertainty in medicine cost resulted in per capita Complete UHCC cost ranges of $223-378, $277-433, and $352-552 per capita for LIC, LMIC and UMIC, respectively. Uncertainty around PiN sizes similarly has a significant effect on per capita Complete UHCC cost: $256-3661, $303-433 and $393-527 per capita for LIC, LMIC and UMIC respectively.

## Discussion

We present the first global, ingredient-based costing model built on the WHO UHCC, combining authoritative epidemiological and demographic data with detailed, service delivery resource estimates. Resource needs and costs are estimated for a comprehensive set of essential health interventions at the level of specific health encounters, enabling analysis that directly supports planning, budgeting and delivery. The model builds directly on a costing tool previously developed and implemented for national benefits package reform in Kyrgyzstan, demonstrating its feasibility and policy relevance in real-world decision-making.(33) As such, the approach provides a structured starting point for national contextualization and use in policymaking.

UHCC Core cost estimates are higher than those reported for the DCP-3 EUHC package. Delivering Core UHCC interventions at 80% coverage is estimated at $2.0 trillion across LMICs (5.7% of GDP), with per capita costs 1.7–2.7 times higher than DCP-3’s original EUHC estimates in LMICs. These differences primarily reflect more comprehensive intervention specifications in the UHCC, rather than differences in disease scope or target coverage. Comparisons with the more restricted DCP-3 HPP further illustrate the substantially lower resource requirements associated with narrower intervention sets.

Estimated Core UHCC costs align closely with WHO projections for achieving SDG3 across LMICs. However, the two approaches differ in purpose and scope. WHO SDG3 costing frames resource needs along a goal-oriented investment pathway and includes population interventions, whereas the UHCC provides implementable descriptions of clinical interventions, specifically designed to support national benefits package design. The detailed resource descriptions now also allow for costing and health system planning.

The UHCC does not constitute a WHO-recommended or minimum UHC package, but is a list of interventions for consideration for inclusion in country-led package development. Further conceptual clarity on the distinction between Core and Additional interventions, explicitly linked to WHO priority-setting guidance, would strengthen the interpretability and policy relevance of UHCC-based cost estimates. At the level of individual interventions, the UHCC incorporates WHO-defined normative standards for service delivery, specifying how interventions are delivered in terms of encounter frequency, inputs and clinical processes.

These service delivery standards should be interpreted as aspirational. For example, the estimated 23 outpatient visits per capita and the medication consumption rates exceed current utilization levels, and reflect universal utilization patterns for, e.g. overweight, hypertension, infectious disease, diabetes, and preventive care currently not attained in many countries worldwide (**Supplement table 4**). This underscores the need for national contextualization of the intervention list and their related standards of care before use in national health policy, through priority-setting, benefits package design and implementation.

### Limitations

This study has several limitations. First, we applied largely static unit costs for drugs, tests, consumables and other high-cost inputs across income groups, reflecting data availability rather than country specific variation. While consistent with previous modelling efforts, this approach does not capture differences arising from trade barriers, taxation, and effects of production and procurement policy. Second, we did not distinguish between public and private provision, despite known differences in cost structures and service delivery patterns.(34) Third, several parameters were held constant across countries, including hours worked per fte, cost of medicine and consumables, procedure duration and operating team size, despite documented variation in actual service delivery. Fourth, our sensitivity analysis incorporated available uncertainty ranges for PiNs, and we constructed variance ranges for medicines and overheads based on literature. We were, however, unable to identify or reconstruct confidence intervals for wages, IVDs, or other products.

These limitations constrain the direct applicability of point estimates to individual countries and underscore the importance of national contextualization. A key strength of the Excel-based model is that its assumptions are transparent and can be readily adapted using country specific data and expert input. We anticipate that this flexibility will help lower barriers to ingredient-based intervention costing and health system modelling in support of evidence-informed EPHS design, implementation, policymaking and advocacy.

## Conclusion

Using a transparent, ingredient-based model built on the WHO UHCC, we estimated the resource requirements associated with normative delivery of 544 essential interventions across 122 LMICs. Delivering Core UHCC interventions at 80% coverage could require approximately $2.0 trillion annually, corresponding to $249, $294, and $363 per capita in low-, lower-middle-, and upper-middle-income countries, respectively. These figures reflect normative service standards and should be interpreted as reference estimates rather than prescriptive spending needs or global targets. In addition, this work provides an adaptable framework to support country-level contextualization and health benefit package design.

## Supporting information

Supplement UHC Costing Model Calculations & Formulas

Table Per Capita UHC Intervention Costs

Dataset Default Costs & Assumptions

## Data Availability

All data produced in the present study are available upon reasonable request to the authors

## Author contributions

*CRediT (Contribution Roles Taxonomy): C: Conceptualization, M: Methodology, I: Investigation, FA: Formal Analysis, W: Writing - First Draft, R: Writing - Revise and Review*

Charlie Nederpelt: C M I FA W

Gerard Abou Jaoude: C M I FA R

Gavin Surgey: M I R

Baktygul Isaeva: I R

Saltanat Zhetibaeva: I R

Han Win Htat: R

Hassan Haghparast-Bidgoli: M I FA R

Rob Baltussen: C M I FA R

## Supplementary Tables and Figures

**Table S1.**
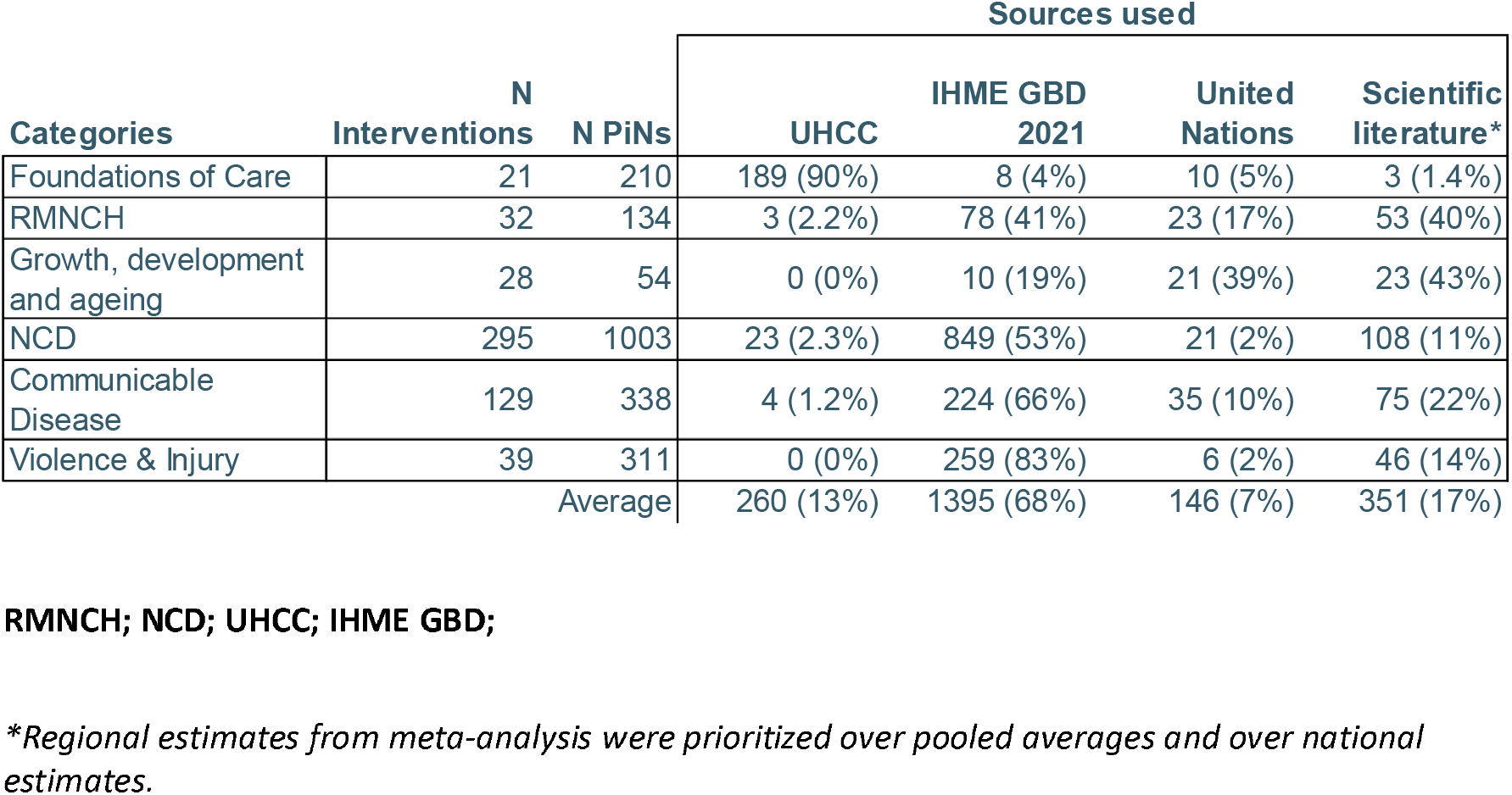
Sources used for population-in-need estimates for healthcare encounters and actions.

**Table S2.** Total estimated cost and proportion of cost components across World Bank income groups.

|  | *All LMICs* |  | *LIC* |  | *LMIC* |  | *UMIC* |  |
| --- | --- | --- | --- | --- | --- | --- | --- | --- |
|  | Cost | % | Cost | % | Cost | % | Cost | % |
| **Total cost (x 1 bln)** | \$2,509 | 100% | \$ 295 | 100% | \$ 995 | 100% | \$1,219 | 100% |
| Health worker | \$ 376 | 15% | \$ 23 | 8% | \$ 146 | 15% | \$ 208 | 17% |
| Medicine | \$1,002 | 40% | \$ 134 | 46% | \$ 392 | 39% | \$ 476 | 39% |
| IVD | \$ 139 | 6% | \$ 24 | 8% | \$ 63 | 6% | \$ 52 | 4% |
| Products | \$ 423 | 17% | \$ 47 | 16% | \$ 170 | 17% | \$ 205 | 17% |
| Overhead | \$ 569 | 23% | \$ 67 | 23% | \$ 224 | 23% | \$ 278 | 23% |

**Figure S1.**
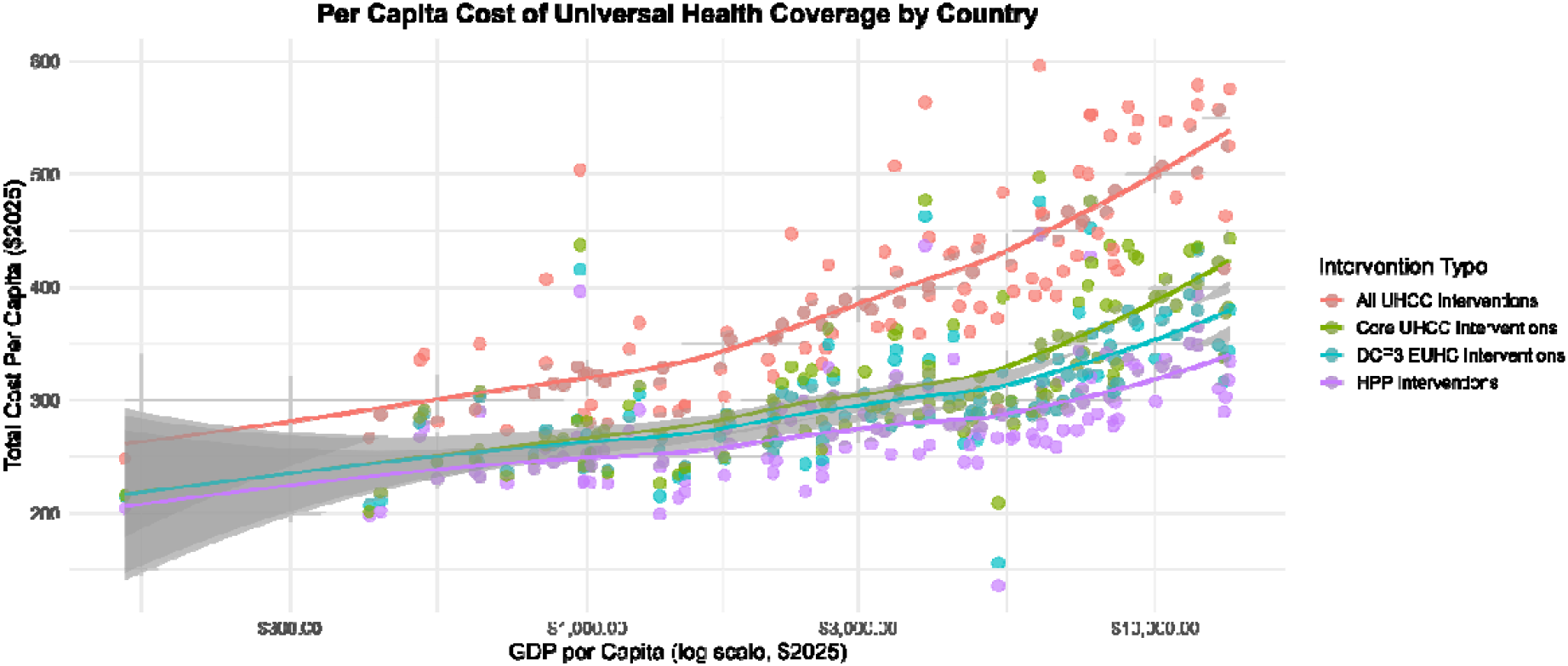
Association between national gross domestica product per capita and per capita cost of four combinations of UHC interventions.

**Figure S2.**
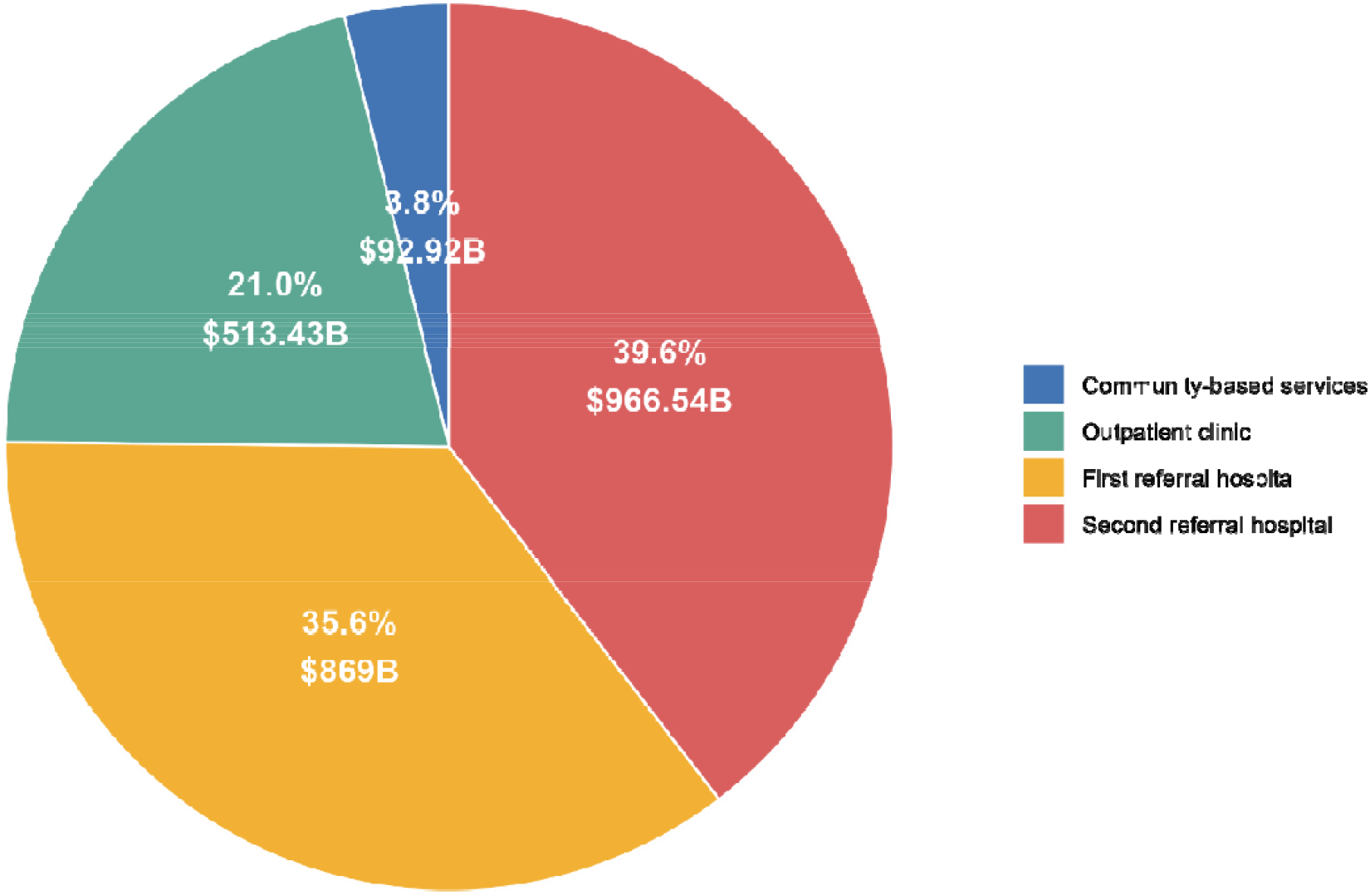
Distribution of cost per delivery channel

**Table S3.**
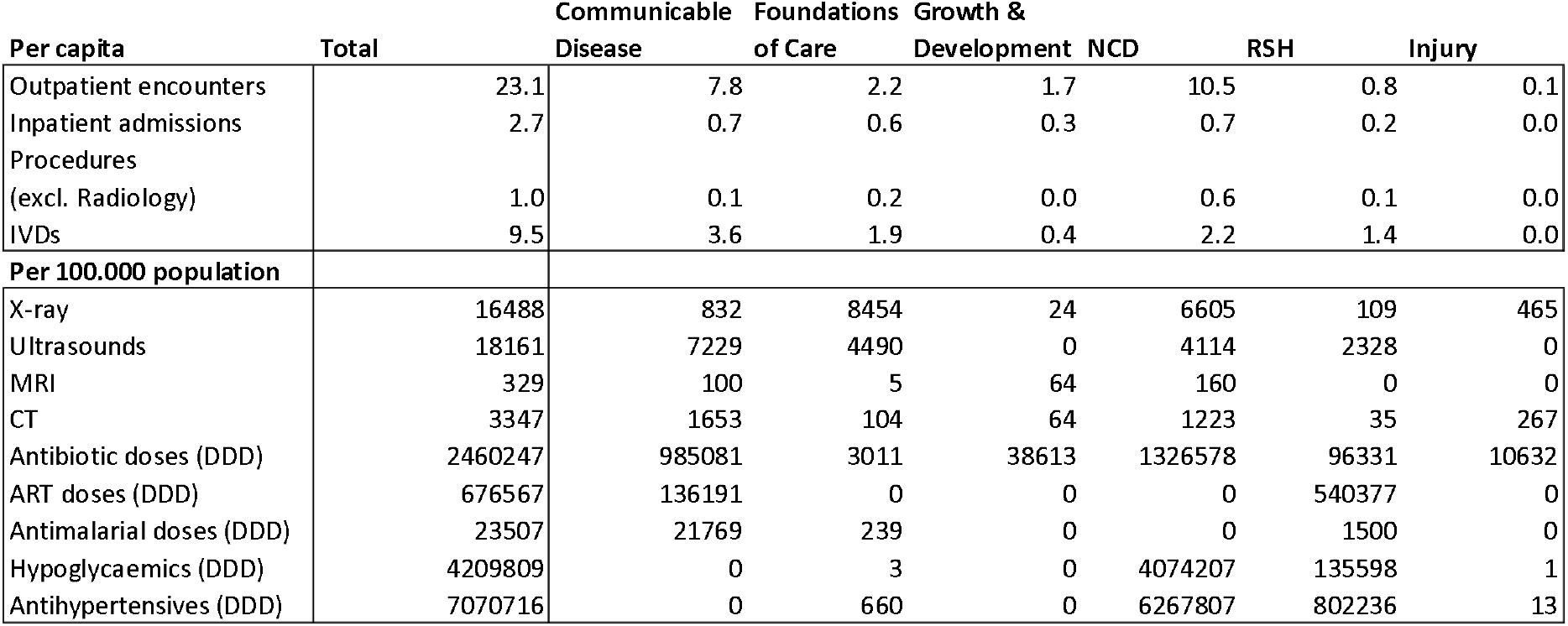
Selected resource use per intervention category across all LMICs.

**Table S4.**
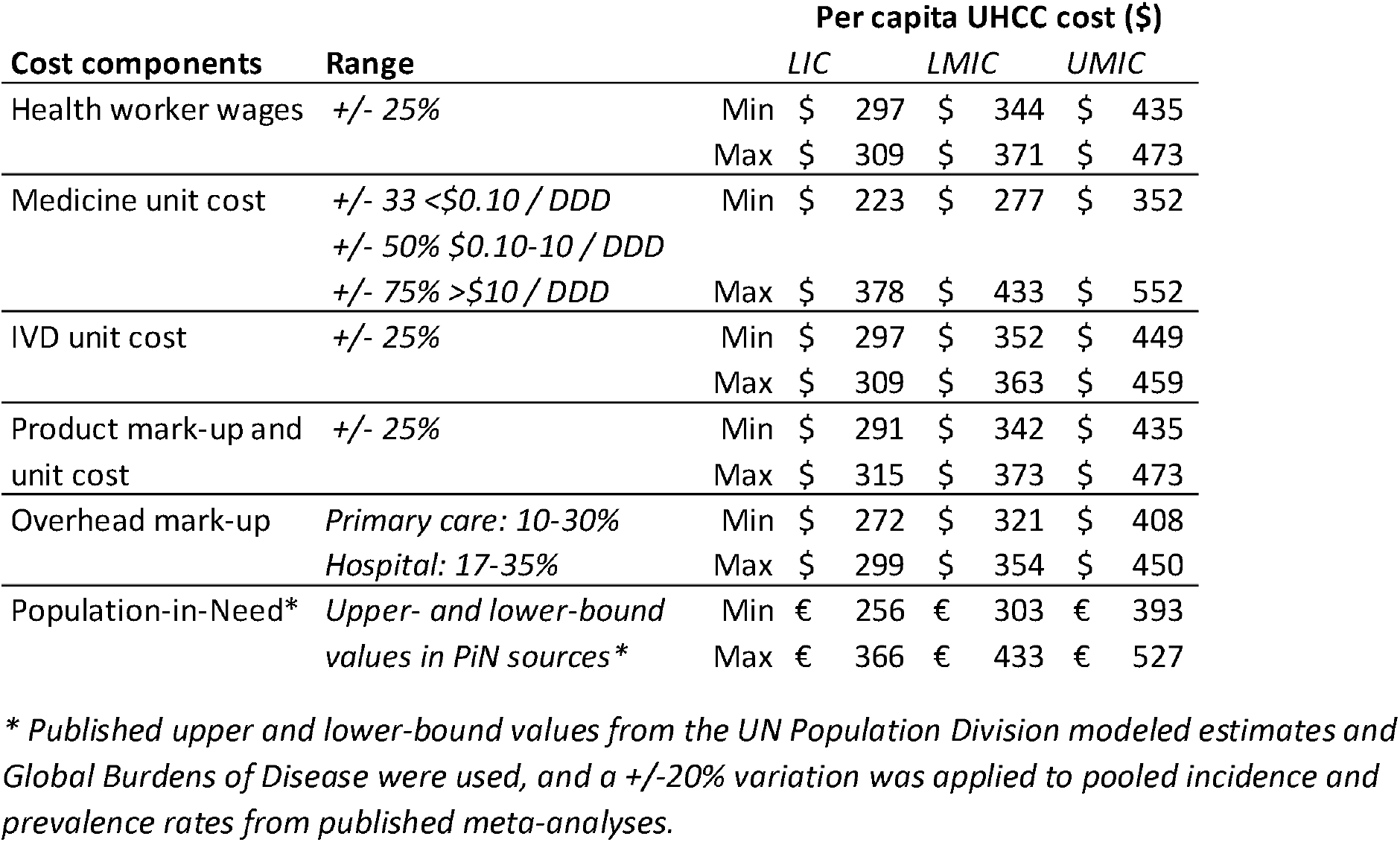
Variation in per capita cost of Complete UHCC interventions based on sensitivity analyses per cost component per World Bank income group.

| Cost components | Range | Per capita UHCC cost (\$) | | | |
| --- | --- | --- | --- | --- | --- |
|  |  |  | LIC | LMIC | UMIC |
| Health worker wages | +/- 25% | Min | \$ 297 | \$ 344 | \$ 435 |
| | | Max | \$ 309 | \$ 371 | \$ 473 |
| Medicine unit cost | +/- 33 <\$0.10 / DDD | Min | \$ 223 | \$ 277 | \$ 352 |
| | +/- 50% \$0.10-10 / DDD | | | | |
| | +/- 75% >\$10 / DDD | Max | \$ 378 | \$ 433 | \$ 552 |
| IVD unit cost | +/- 25% | Min | \$ 297 | \$ 352 | \$ 449 |
| | | Max | \$ 309 | \$ 363 | \$ 459 |
| Product mark-up and unit cost | +/- 25% | Min | \$ 291 | \$ 342 | \$ 435 |
| | | Max | \$ 315 | \$ 373 | \$ 473 |
| Overhead mark-up | Primary care: 10-30% | Min | \$ 272 | \$ 321 | \$ 408 |
| | Hospital: 17-35% | Max | \$ 299 | \$ 354 | \$ 450 |
| Population-in-Need* | Upper- and lower-bound | Min | € 256 | € 303 | € 393 |
|  | values in PiN sources* | Max | € 366 | € 433 | € 527 |

\* Published upper and lower-bound values from the UN Population Division modeled estimates and Global Burdens of Disease were used, and a +/-20% variation was applied to pooled incidence and prevalence rates from published meta-analyses.

## Supplement 3 Robustness analysis

We modeled normative human resource requirements of 6.15 fte/1,000 population, 18-43% lower than WHO (7.6 and 9.0/1,000 for LIC and LMIC/UMIC) and GBD projections (10.9/1,000 population) based on actual service delivery.(31,32) We find significantly higher (4x) utilization of outpatient visits than previous studies at 23 vs 5.4-5.9 visits/capita.(31,35) UHCC includes several high-utilization interventions for prevention, diagnosis and treatment of communicable diseases (8/capita) and NCDs (11/capita) (**Table S2**). This includes interventions not commonly delivered universally even in high-income countries, such as overweight and obesity (2.6/capita), depression (1.0/capita), and oral health (1.0/capita). Our estimate of 2.7 admission days p.p.p.y. is likely 3-6x higher than previous estimates, including IHME’s 0.10 (duration unspecified) and Eurostat 0.13-0.16 admissions p.p.p.y. with 4.5-9.7 avg. length-of-stay of.(35,36)

Compared to a proposed minimum surgery rate of 0.05/capita, we estimate non-radiological procedure utilization at 1/capita, but UHCC includes procedures like airway management, biopsies, closed repositions, casting, etc.(37) Radiography capacity is recommended at 1 X-ray and 1 ultrasound machine per 50,000 population, which is realistic for our estimates of 8,222 X-rays and 9.293 ultrasounds per 50,000 people p.y., or 23 and 25 per day.(38) Literature on IVD utilization in LMICs was not found, but our estimate of 10 IVDs p.p.p.y. matches England and Wales (14 p.p.p.y.) and Ontario, Canada (3.7 p.p.p.y.).(39,40)

Published data on drug consumption reflect both under- and overuse. Estimates of UHCC medication consumption are 4 times higher for antibiotics (23 vs 5.1 DDD p.p.p.y. in LMICs), 3 times higher for antihypertensives (67 vs 21 DDD p.p.p.y.), and 9 times higher for hypoglycaemics (37 vs 4 DDD p.p.p.y.).(41–43) We furthermore estimate 43.6 billion annual ART doses, corresponding to ca. 1100 DDDs per year for the 40.8 million PLHIV requiring ART in the latest UNAIDS report.(44) We estimate an annual consumption of 1.5 billion DDDs for both chemoprophylaxis and the 263 million incident cases in 2024.(45)

Cost comparisons have common limitations, including differences in intervention definition, detail and aggregation level, normative versus actual delivery, and costing scope. For drug-susceptible TB, we estimated a mean $367, $474, $852 for DS-TB in LIC, LMIC and UMIC respectively; and $6069, $6110, and $6142, for drug-resistant TB. This matches well to inflation-adjusted published estimates of $361, $371, and $1142 for DS-TB in LIC, LMIC and UMIC respectively, and to 2023 WHO estimates of median $783 for DS-TB, but less closely to the $4877 MDR-TB estimate.(46,47) We estimated a mean cost for testing and treatment across LMICs of $18 for uncomplicated and $394 for severe malaria (including parenteral medication, transfusion and oxygen therapy) compared to inflation published adjusted estimates of $20 and $122.(48) Annual cost of diagnosing and treating diabetes, including complications, were estimated at $93 per confirmed case (medication costs $60/case), which falls within the reported range for LMICs of $26-57 for medication, and 29-990$ for visits and admissions, although reported costs vary significantly.(49) ART costs per incident case were estimated at a mean $583/case, $406 of which towards medication, much lower than inflation-adjusted published drug costs of $750, $690, $1011 across LIC, LMIC and UMIC respectively.(50)

