## Supplement UHC Costing Model Calculations & Formulas for "Resource need and cost estimates for universal health coverage across 122 countries using the WHO UHC Compendium"

**Supplement:** Costing tool calculations summary

Calculations used to estimate costs per intervention in the UHCC Costing Tool are summarized below. Intervention costs are summed across relevant platforms.

**Notation**

- Service delivery platforms: $p\in\{\text{COM}, \text{OPT}, \text{1RL}, \text{2RL}\}$
  - COM = Community-based
  - OPT = Outpatient/clinic
  - PRE = Pre-hospital level
  - 1RL = First referral level
  - 2RL = Second referral level
- Let:
  - $U_{p}$= Percentage Utilisation Distribution for platform $p$
  - $N$= Annual number of visits or inpatient days
  - $D$= Duration of inpatient stay in days
  - $P_{\text{need}}$= Percentage of population in need
  - $C_{\text{visit},p}$= Cost per visit or inpatient day at platform $p$, based on the type of visit or inpatient stay
  - $W$= Health worker wage per minute
  - $t_{\text{proc}}$= Procedure duration in minutes
  - $s_{\text{action}}$= Percentage share of action
  - $s_{\text{HP}}$= Percentage share of health product
  - $C_{\text{med}}$= Cost per medicine unit, based on route of administration and dosage form
  - $d_{\text{dose}}$= Medicine doses per day
  - $T_{\text{days}}$= Annual number of treatment days
  - $C_{\text{IVD}}$= Cost per diagnostic/IVD unit, based on the type of diagnostic/IVD
  - $N_{\text{IVD}}$= Annual number of diagnostic/IVD units required
  - $C_{\text{Adj}}$= Cost per treatment adjunct unit, based on the type of treatment adjunct
  - $N_{\text{Adj}}$= Annual number of treatment adjunct units required
  - $u_{\text{cons},p}$= Uptick for consumables at platform $p$
  - $u_{\text{over},p}$= Uptick for overheads at platform $p$
  - $C_{\text{excl}}$= Costs excluding treatment adjuncts, consumables and overheads
  - $N_{\text{pin}}$= Number of people in need
  - $U_{acc}$= Current or target percentage of people in need accessing the service
  - $M$= Multimorbidity factor equal to half the ratio between the pooled prevalence of chronic multimorbidity and morbidity in LMICs

**1) Visit / Inpatient Stay Direct Labour Component**

$$C_{\text{visit}}=\sum_{p} (P_{\text{need}}\times C_{\text{visit},p}\times N\text{ or }D\times U_{p})$$

**2) Procedures Direct Labour Component**

$$C_{\text{proc}}=\sum_{p} (t_{\text{proc}}\times W\times s_{\text{action}}\times U_{p})$$

**3) Medicines**

$$C_{\text{med}}=\sum_{p} (C_{\text{med}}\times s_{\text{action}}\times s_{\text{HP}}\times d_{\text{dose}}\times T_{\text{days}}\times U_{p})$$

**4) Diagnostics / IVD**

$$C_{\text{IVD}}=\sum_{p} (C_{\text{IVD}}\times s_{\text{action}} \times N_{\text{IVD}}\times U_{p})$$

**5) Treatment Adjuncts**

$$C_{\text{Adj}}=\sum_{p} (C_{\text{Adj}}\times s_{\text{action}} \times N_{\text{Adj}}\times U_{p})$$

**6) Consumables**

$$C_{\text{cons}}=\sum_{p} (u_{\text{cons},p}\times C_{\text{excl}}\times U_{p})$$

**7) Overheads**

$$C_{\text{over}}=\sum_{p} (u_{\text{over},p}\times C_{\text{excl}}\times U_{p})$$

**8) Total Service Cost per Service User**

$$C_{\text{user}}=C_{\text{visit}}\times M+C_{\text{proc}}+C_{\text{med}}+C_{\text{IVD}}+{C_{\text{Adj}}+ C}_{\text{cons}}+C_{\text{over}}$$

**9) Total Service Cost**

$$C_{\text{total}}=C_{\text{user}}\times N_{\text{pin}}\times U_{acc}$$
